# A protocol for a pilot randomized controlled trial of guided self-help Behavioral Activation intervention for geriatric depression

**DOI:** 10.1101/2020.02.03.20020156

**Authors:** Xiaoxia Wang, Xiaoyan Zhou, Hui Yang

## Abstract

**Background:** Aging is a social concern. Despite increased incidence of depression in Chinese older populations, they are often misdiagnosed and undertreated, which pose a challenge to our health care system. Depressed older adults often suffer from lack of motivation. Evidence-based guided self-help behavioral activation treatment is effective in reducing anhedonia and amotivation symptoms, however the efficacy in in elderly depressed patients are unknown yet. So the current study aim to pilot the self-help guided intervention for the treatment of depression in elderly patients.

**Methods/design:** A pilot randomized controlled trial with 30 elderly inpatients with major depressive disorder will be conducted. Participants attending clinical psychological clinics at Mental Health Center of Chongqing will be randomized to receive guided self-help BA intervention or to a 6-week waiting list control. Participants in the treatment group will receive 6 sessions of guided self-help BA intervention delivered over the telephone. The waiting list control group will receive the intervention after a waiting period of 6 weeks. Exclusion criteria are limited to those at significant risk of harm to self or others, the presence of primary mental health disorder other than depression or intellectual disability at a level meaning potential participants would be unable to access the intervention. The study has ethical approval from the Chinese Research Ethics Committee. Our hypothesis is that guided self-help BA will effective when compared with the waiting-list intervention.

**Results:** Effects of the treatment were observed on three outcomes domains: (1) clinical outcomes (symptom severity, recovery rates); (2) process variables (patient satisfaction, attendance, dropout); and (3) economic outcomes (cost and resource use). We also examine mediators of outcomes in terms of patient variables (behavioral activation/inhibition motivation). Results will be disseminated to patients, the wider public, clinicians and researchers through publication in journals and presentation at conferences.

**Discussion:** This is the first study to investigate guided self-help interventions for China geriatric depressed patients, a group which is currently under-represented in mental health research. The intervention is modular and adapted from an empirically supported behavioral activation treatment for depression (BATD). The generalizability and broad inclusion criteria are strengths but may also lead to some weaknesses.

## Introduction

With the increase in life expectancy and low fertility rate, rapid aging has become a social concern. According to the data from China’s 2010 population census, the percentage of the elderly population aged 65 years and older (14.51%) is higher in Chongqing than in other provinces in China. Chongqing is among the provinces with the most rapid aging (李俊明, 2016). The elderly population aged over 65 years and older in Chongqing in 2020 is predicted to grow to approximately 3.93 million (李苗, 2016). With the increase in the aging population, geriatric depression has become the most important public health issue in the elderly population and can significantly increase the total cost of medical services (Unutzer et al.,1997). Geriatric depression specifically refers to primary depression that first occurs after the age of 60 and cannot be explained by physical symptoms or other organic diseases (徐杰,2014). The prevalence rate of depressive symptoms in the central and western regions of China (33. 7%) is significantly higher than that in the eastern region (19.1%) (张玲, 徐勇, & 聂宏伟,2011). Among elderly people investigated in Chongqing, 24.3% had different degrees of depressive symptoms (Giri, Chen, Yu, & Lu,2016).

Currently, depression in older adults is treated mainly in primary care or home-based setting (Nguyen & Vu,2013) and often inadequately treated (Ghio, Vaggi, Amore, Ferrannini, & Natta,2014). Among the investigated sample, only one-third of depression patients will receive appropriate treatment, 25%-33% of untreated depression patients will continue to have symptoms, and the suicide rate will reach 15% (薛海波, 2004). The most common treatment modalities for MDD are antidepressant drugs and psychological interventions (or a combination of both) (Clignet, van Meijel, van Straten, & Cuijpers,2012). Evidence-based treatment of geriatric depression mostly focus on pharmacological treatment (Ghio et al.,2014). Recent reviews support the efficacy of psychosocial interventions in the acute treatment of geriatric depression (Cuijpers, Karyotaki, Pot, Park, & Reynolds,2014;Kiosses, Leon, & Areán,2011; 王锋, 吴红梅, 黄昶荃, 卢振产, & 董碧蓉,2008). Behavioral therapy in depressed older adults appears to have comparable effectiveness with alternative psychotherapies (Samad, Brealey, & Gilbody,2011), with its efficacy not lower than that of cognitive behavioral therapy (Richards et al.,2016). Notably, there are major gaps between the efficacy of treatments in controlled clinical trials and the effectiveness of treatment in a clinical setting. Behavioral therapy is a briefer, simpler intervention which might benefit older adults with a low educational level or suffer from cognitive impairment.

The monitoring and scheduling of activities are common components in all treatment protocols used in randomized studies of the BA approach (Pasterfield et al.,2014). Behavioral activation therapy increases the positive reinforcement (such as obtaining a sense of pleasure and control) following antidepressant behaviors (such as completion of scheduled activity, active social engagement) which then improves patients’ depression symptoms (Takagaki et al.,2016). Furthermore, brain functional magnetic resonance imaging study indicated that behavioral activation therapy can improve depression symptoms while increasing the reward brain system (dorsal striatum, orbital frontal gyrus) activation to rewards (Dichter et al.,2009). Activity scheduling as effective component of BA showed significant effect size when compared with control condition (Cuijpers, van Straten, & Warmerdam,2007). The development of enjoyable leisure activities have an adequate level of evidence?(Dirmaier et al.,2012). However, the application of BA to late-life depressed patients has been underexplored.

As the tele-mental health (TMH) service mode increasingly matures, guided self-help treatment can be performed regularly under the guidance of consultants using smart cell-phones and the internet. According to the NICE-guideline, guided self-help intervention delivered without human support or guidance is assigned the evidence degree II (Dirmaier et al.,2012). Compared to drug treatment, patients are more inclined to receive guided self-help therapy and psychotherapy (Hanson, Webb, Sheeran, & Turpin,2015). A meta-analysis confirmed similar effects between guided self-help CBT and face-to-face CBT for depressive disorders in primary care (Linde et al.,2015). However, there is still limited report on the implementation of guided self-help BA interventions for geriatric depression. Only one study using BA bibliotherapy (Addis and Martell’s Overcoming depression) one step at a time as a stand-alone treatment over a 4-week period, which showed decreased symptoms on a clinician-rated measure of depressive symptoms (HAMD) (Moss, Scogin, Di Napoli, & Presnell,2012).

Therefore, a guided self-help BA intervention is adopted to implement psychological education about depression and behavioral activation plans for geriatric depression patients. The behavioral activation intervention application previously developed by our research group (unpublished) is used for monitoring and scheduling of pleasurable activities. The geriatric depression scale combined with the behavioral activation inhibition scale (BIS) is used to investigate the effect of improvement of geriatric depression symptoms using the guided self-help behavioral activation intervention.

### Participants

A total of 60 subjects who conformed to the diagnostic criteria of geriatric depression were randomly selected. Subjects were randomly divided into the study group and the control group, with 30 individuals in each group. The general data (gender, age, education level, marital status, occupation, residence, income, and disease course) between the two groups did not demonstrate significant differences.

[Inclusion criteria] (1) Depression patients conformed to the International Classification of Diseases (ICD)-10 diagnostic criteria; (2) patients had resided in the area for more than 6 months; (3) patients had barrier-free communication and were able to complete questionnaires independently or with assistance; (4) patients aged between 60-70 years, with an education level of junior high school or above, and frequent use of smartphones (could receive and check text messages and perform psychological assessment using a simple visualized scale on phones).

[Exclusion criteria] (1) Patients with severe physical illness and organic brain diseases; (2) patients with confirmed schizophrenia spectrum, psychoactive substance use disorder, and/or mental retardation; (3) patients with bipolar disorder; (4) patients with a recent history of severe infection and fever; (5) patients with severe suicidal tendency; (6) patients with other conditions that were not suitable for inclusion after evaluation by researchers.

### Study design

The study group and the control group were both given selective serotonin reuptake inhibitor (SSRI) treatment, conventional care, and health education. However, behavioral activation intervention was added only to the study group. For the study group, the program was formulated based on the short-term behavioral activation therapy program of Lejuez et al. (Lejuez, Hopko, Acierno, Daughters, & Pagoto,2011) combined with the hospital condition. This treatment is performed for 40-50 min each time and twice each week for a total of 3 weeks (Fig 2). Except for the group intervention, which provides uniform guidance on the behavioral activation program for geriatric depression patients, the behavioral intervention software we developed was used to perform depression symptom assessment and monitor the completion condition of behavioral activation tasks during the treatment course and after completion of the treatment.

**Fig 2.**
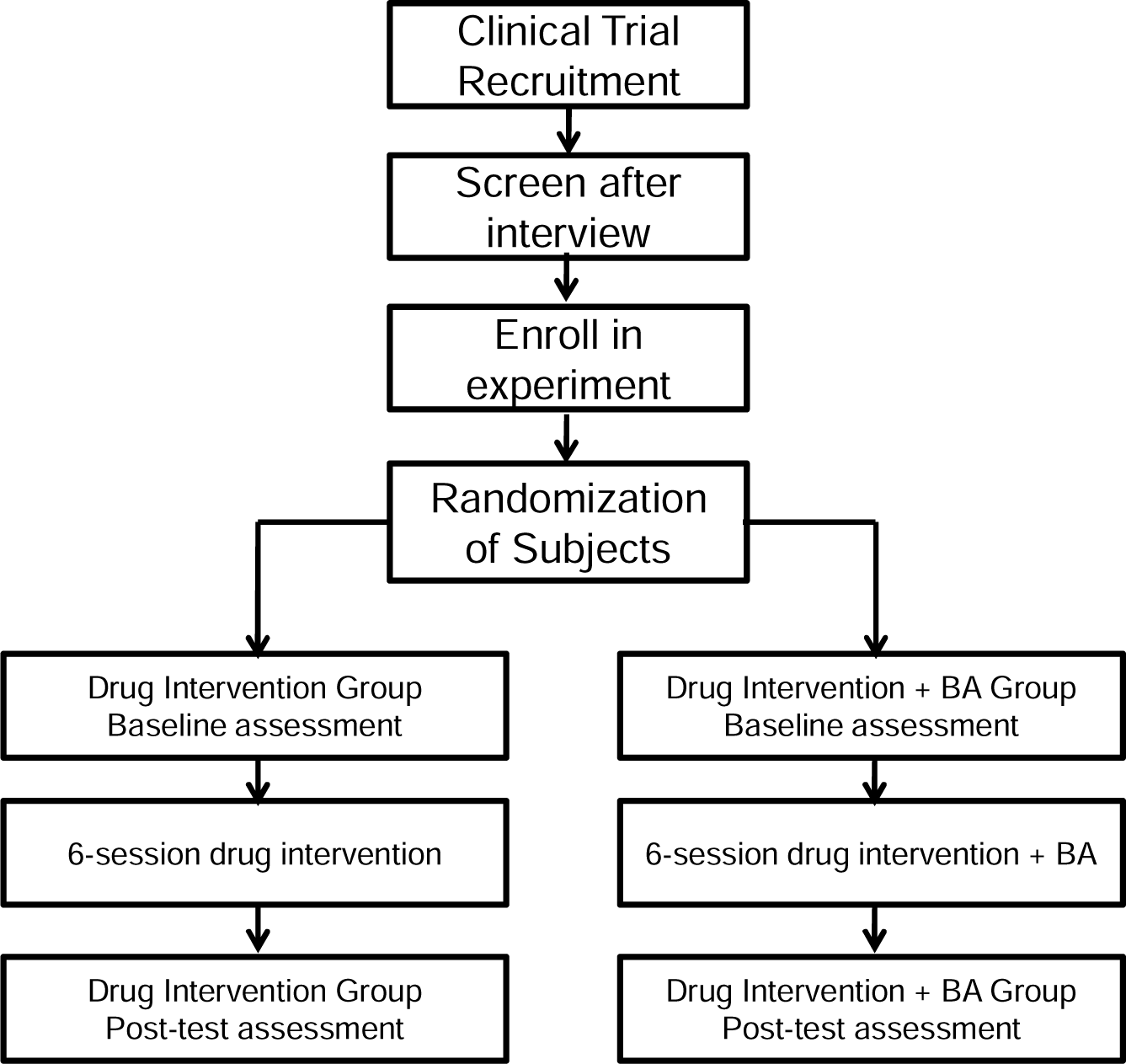
Study design of guided self-help behavioral activation intervention for geriatric depression

### Treatment sessions

There are six treatment sessions. Afterward, monitoring of the depression condition and the behavioral activation task completion condition is performed using the depression behavioral activation intervention software we developed. (1) The first session provides psychological education about depression. The treatment concept and the status and importance of daily monitoring in the treatment process are introduced to visitors. (2) The second session involves a review of the previous treatment course to introduce the value and concepts of the activity plan. In different life domains (such as family relationships, learning and work, leisure activities, and hobbies and interests), visitors and therapists are inspired through suggestion and writing tasks to list a series of activities that they feel are valuable and enjoyable. The activity schedule is listed together with therapists to allow visitors to obtain maximum positive reinforcement in life. (3) During the third and the fifth session, the treatment concept is constantly reviewed and revised. Visitors are asked to make an activity list using the activity plan. (4) The sixth session is used to discuss the completion of the treatment, evaluate treatment progress, and teach visitors to prevent recurrence using the knowledge and skills learned from treatment (Luxton et al.,2014) (Table 1). After group intervention, the above program combined with the behavioral activation intervention software we developed can assist individuals with the self-learning of psychological knowledge of depression and behavioral activation interventions, establishment of an activity schedule, and the monitoring of daily emotional changes. Each session uses the self-rating scale (including two methods: software self-help assessment and paper questionnaire) to monitor depression symptoms and motivation changes in patients.

**Table 1.**
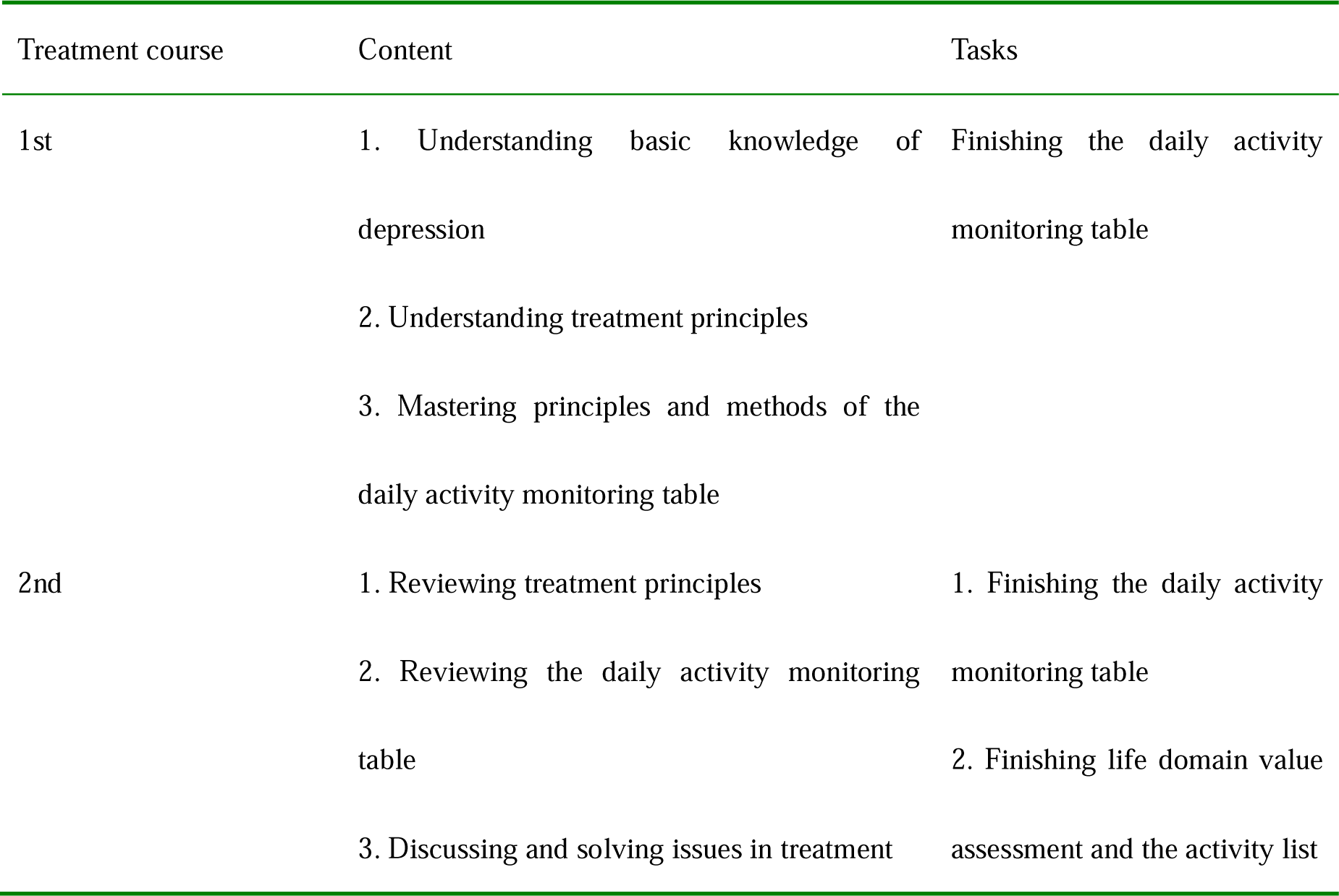
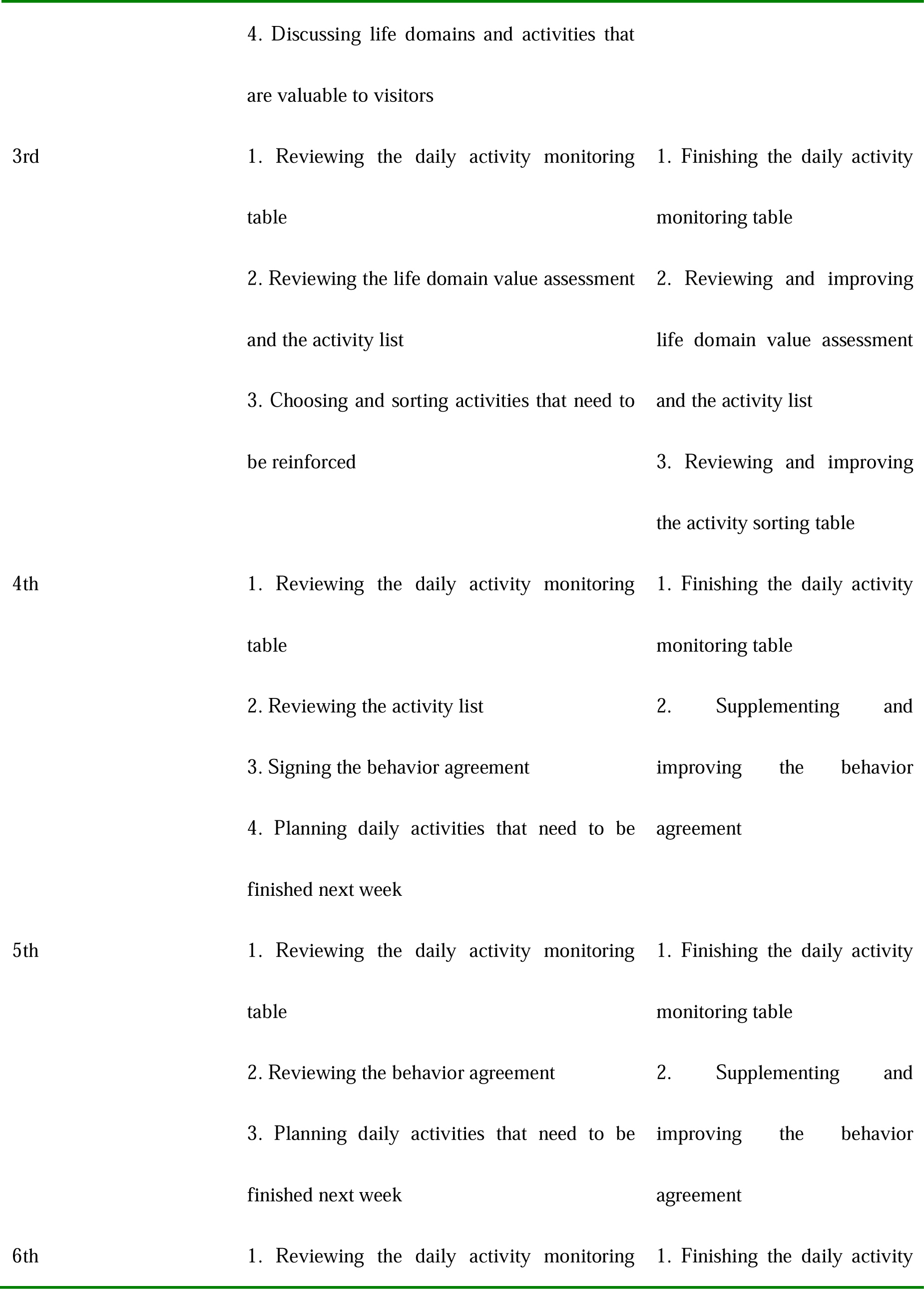
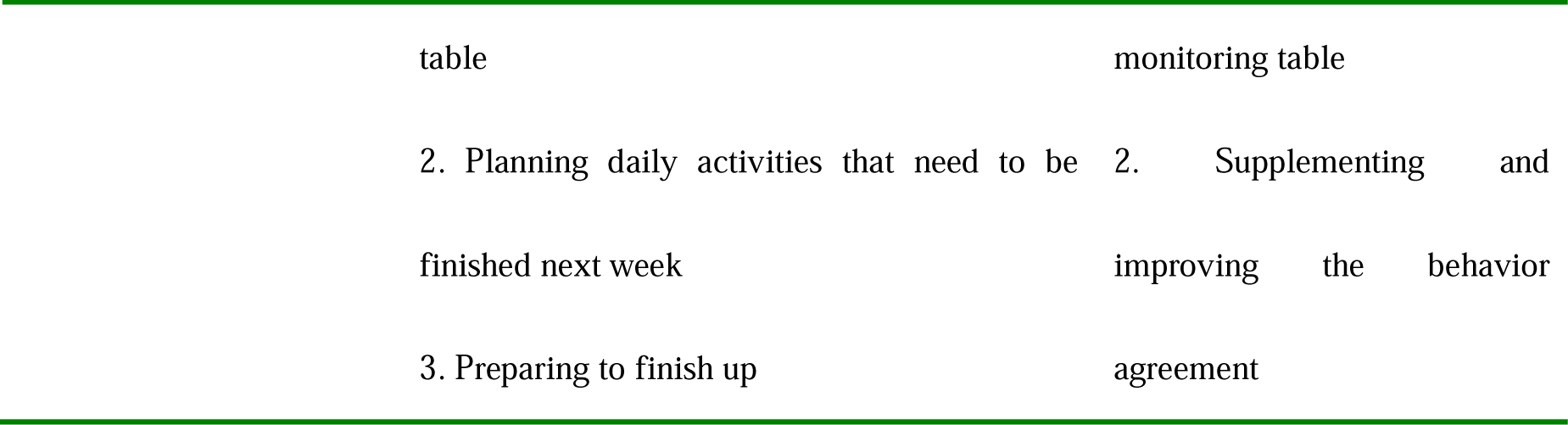
Establishment of the treatment courses of behavioral activation therapy

### Quality control

Study subjects were selected strictly according to the inclusion and exclusion criteria. (2) Medical care staff with patience, technique, and strong responsibility were chosen for performing guidance. They received uniform, rigorous training before performing guidance. (3) Attention should be paid to communication skills and protection of patients’ privacy. (4) The data input and review system was established to ensure the accuracy of data input and analysis.

### Health Outcomes

(1) The Geriatric Depression Scale (GDS). This scale is a self-rating depression scale specifically developed for elderly people. This instrument includes a total of 30 items. The statistical indicator of this scale is the total score. A total score of 0-10 points indicates no depression symptoms, 11-20 points indicates mild depression symptoms, 21-25 points indicates moderate depression symptoms, and 26-30 points indicates severe depression symptoms.

(2) The Behavioral Inhibition System and Behavioral Activation System Scale (BIS/BAS Scale). This scale was developed by Carver and White in 1994 based on the theory by Gray. Li et al. translated it into Chinese in 2008. This scale has 20 items and is divided into two systems: the behavioral inhibition system and the behavioral activation system (including three dimensions: reward response, drive, and fun seeking). This scale has 1 to 4 points, ranging from “completely agree” to “completely disagree.” The Cronbach’s α of all dimensions ranges from 0.66-0.76. The reliability measured after 2 months is 0.59∼0.69.

### Efficacy Outcome

After follow-up for 6 months, the recurrence rate was evaluated using the Clinical Global Impression Scale (CCG-GL). The Treatment Emergent Symptom Scale (TESS) was used to evaluate side effects.

### Statistical analysis

Statistical examination methods such as independent and paired sample t tests were performed using the SPSS 22.0 software (SPSS, Inc., Chicago, IL, USA). The effect of improvement in geriatric depression symptoms using behavioral activation intervention was investigated. Amos 17.0 software (SPSS, Inc., Chicago, IL, USA), combined with the mediation effect model, was used to investigate the mediating effect of the behavioral activation inhibition level in the improvement of geriatric depression symptoms using the behavioral activation intervention. The mediation effect model and the path analysis statistical method are used to investigate the mediating mechanism using the behavioral activation/inhibition motivation level as the treatment effect of the guided self-help behavioral activation intervention.

### Ethics and dissemination

#### Ethical approval

This study has been reviewed and approved by the Research Ethics Committee of Mental Health Center of Chongqing. All participants will give verbal and written informed consent before inclusion and randomization. The study has been registered with the Current Controlled Trial (ChiCTR1900026066).

#### Informed consent

The results of this study are for scientific purposes only. All information collected during the study is confidential and will be protected in accordance with the law. The participants’ name and identity will not be disclosed. The study data will be kept confidential and will be kept in the hospital. Only the medical staff, ethics committee and national health administration involved in the study will have access to the participants’ confidential information during the necessary supervision and inspection.

#### Ethical and safety considerations

The study involved obtaining the subjects’ basic information or psychological tests mainly through questions and questionnaires, with no risk of physical damage to the subjects. To prevent the possibility that the assessment and intervention may cause the participants to be distressed, the clinically well-trained psychiatrist with over 10 years of professional experience will be prepared to intervene once the participants reported their distressed feelings.

## Discussion

This is the first study to investigate guided self-help behavioral activation interventions for Geriatric Depression — a mental condition which is currently underrepresented in elderly patients. The intervention is modular and adapted from Lejuez’ s manualized behavioral activation treatment for depression (BATD) (Lejuez, Hopko, Acierno, Daughters, & Pagoto,2011). The weakness of the study is generalizability which is limited in the inpatients of Geriatric Depression, which is commonly comorbid with physical illnesses. In addition, this initial study will be based in Chongqing, the western city of China, which experiences the most rapid aging speed.

However, it also has a number of strengths, including random allocation and blinding of study assessors, enhancing internal validity of the results. In addition, the moderators of health outcome measures in BA focused on behavioral activation/inhibition motivation, which is underrepresented in previous study on BA intervention. We also focused on the clinical efficacy as well as effectiveness of guided self-help intervention in the context of the inpatient service of a psychiatric hospital.

## Data Availability

The data of the clinical trial is available upon request to the corresponding author.

## Acknowledgements

This research was financially supported by the Youth Cultivation Foundation of Medical Science in Army Medical University (2016XPY08), the National Youth Cultivation Foundation of Military Medical Science (17QNP002), and the Chongqing Social Science Planning Project (2017QNSH21).

## Notes

### Competing Interest Statement

The authors have declared no competing interest.

### Clinical Trial

ChiCTR1900026066

